# Projected HIV and Bacterial STI Incidence Following COVID-Related Sexual Distancing and Clinical Service Interruption

**DOI:** 10.1101/2020.09.30.20204529

**Authors:** Samuel M. Jenness, Adrien Le Guillou, Christina Chandra, Laura M. Mann, Travis Sanchez, Daniel Westreich, Julia L. Marcus

## Abstract

**Background:** The global COVID-19 pandemic has the potential to indirectly impact the transmission dynamics and prevention of HIV and other sexually transmitted infections (STI). Studies have already documented reductions in sexual activity (“sexual distancing”) and interruptions in HIV/STI services, but it is unknown what combined impact these two forces will have on HIV/STI epidemic trajectories.

**Methods:** We adapted a network-based model of co-circulating HIV, gonorrhea, and chlamydia for a population of approximately 103,000 men who have sex with men (MSM) in the Atlanta area. Model scenarios varied the timing, overlap, and relative extent of COVID-related sexual distancing in casual and one-time partnership networks and service interruption within four service categories (HIV screening, HIV PrEP, HIV ART, and STI treatment).

**Results:** A 50% relative decrease in sexual partnerships and interruption of all clinical services, both lasting 18 months, would generally offset each other for HIV (total 5-year population impact for Atlanta MSM: −227 cases), but have net protective effect for STIs (−23,800 cases). Greater relative reductions and longer durations of service interruption would increase HIV and STI incidence, while greater relative reductions and longer durations of sexual distancing would decrease incidence of both. If distancing lasted only 3 months but service interruption lasted 18 months, the total 5-year population impact would be an additional 890 HIV cases and 57,500 STI cases.

**Conclusions:** The counterbalancing impact of sexual distancing and clinical service interruption depends on the infection and the extent and durability of these COVID-related changes. If sexual behavior rebounds while service interruption persists, we project an excess of hundreds of HIV cases and thousands of STI cases just among Atlanta MSM over the next 5 years. Immediate action to limit the impact of service interruptions is needed to address the indirect effects of the global COVID pandemic on the HIV/STI epidemic.

## INTRODUCTION

The 2019 novel coronavirus (COVID-19) global pandemic has directly resulted in substantial morbidity and mortality, but has also indirectly impacted the transmission of other infectious diseases.^1^ For HIV and other sexually transmitted infections (STIs), behavioral responses to COVID-19 have included major reductions in social contacts that also entailed reductions in sexual activity (“sexual distancing”).^2–4^ The pandemic also has interrupted the provision of clinical services for HIV/STIs.^5^ One critical question is how these two phenomena — distancing that could decrease HIV/STI transmission, and service interruption that could increase transmission — will impact the overall incidence of HIV and STIs immediately and in the post-COVID era.

In the United States, men who have sex with men (MSM) are a key population for HIV/STI prevention.^6^ COVID-19 has already resulted in sexual distancing for MSM.^4,7–9^ Behavioral changes that began during COVID-related restrictions (March 2020) have included reductions in the number of sexual partners (40–60% of MSM) and lower frequency of sexual activity within partnerships (20%). While some social activities have already rebounded in some areas,^10^ resumption back to “normal” levels may be delayed until the widespread distribution of a SARS-CoV-2 vaccine. Use of clinical HIV/STI services has also declined among MSM in the U.S. Categories of reduced services include HIV/STI diagnostic screening (25–85%), use of HIV preexposure prophylaxis (PrEP) (20–72%), and retention in HIV care (25%–45%).^7,9^ Providers have partially addressed these interruptions by replacing in-person clinical visits with telehealth services,^11^ but these tools may be less available to people with the greatest need.^12^ Local health departments have also redeployed STI services towards COVID-19 contact tracing efforts.^13^ It is unknown what the immediate and longer-term impact of these service disruptions will be for uniquely for HIV versus STI incidence.

Parallel reductions in sexual behavior and HIV/STI screening rates make it challenging to understand whether any declines in reported case data reflect true changes in disease incidence or gaps in disease surveillance. Transmission modeling can disentangle these two explanations. In this study, we used a stochastic network-based transmission model to project the impact of sexual distancing versus HIV/STI service interruptions driven by COVID-19. We evaluated how changes to the sexual partnership networks among MSM in Atlanta may reduce the disease incidence in a model that represents the cocirculation of HIV, gonorrhea, and chlamydia. We also explored how COVID-related service disruptions in four categories (HIV screening, HIV PrEP, HIV treatment, and STI treatment) could increase the incidence of these infections.

## METHODS

### Study Design

This model of HIV/STI transmission dynamics for U.S. MSM was built with the *EpiModel* platform,^14^ which simulates epidemics over dynamic contact networks using temporal exponential random graph models (TERGMs).^15^ This builds on our previous applied HIV/STI modeling of co-circulating HIV, *Neisseria gonorrhoeae* (NG), and *Chlamydia trachomatis* (CT) among MSM.^16^ For this study, we implemented time points for the start and end of COVID-related sexual distancing and clinical service interruptions. Full methodological details are provided in the **Supplemental Appendix**.

Our model represented main, casual, and one-time sexual partnerships for Black, Hispanic, and white/other MSM, aged 15 to 65, in Atlanta. The starting network size in the model simulations was 10,000 MSM, which could stochastically increase or decrease over time based on arrival (sexual debut) and departure (mortality or sexual cessation).

### HIV/STI Epidemic Model

The epidemic model consisted of 5 main components: 1) statistical network models (TERGMs) to generate dynamic sexual partnerships; 2) statistical models to predict behavior within partnerships; 3) simulation of pathogen transmission across active partnerships; 4) simulation of disease progression and other natural history features; and 5) simulation of prevention and treatment service engagement.

To fit the network models, we used data from ARTnet, a web-based egocentric network study conducted in 2017–2019 of MSM in the US.^17^ Parameters were weighted by census-based race/ethnicity and age distributions to account for ARTnet sampling biases. Multivariate predictors of partnership formation included partnership type, degree distributions by partnership types, heterogeneity in network degree, assortative mixing by race/ethnicity and age, and mixing by sexual position. Generalized linear m models were then fit to ARTnet to predict the frequency of acts and the probability of condom use as a multivariate function of race/ethnicity, age, diagnosed HIV status, and partnership type and duration.

MSM could be screened for HIV and initiate anti-retroviral therapy (ART), which would lower their HIV viral load (VL) and increase their longevity. MSM progressed through HIV disease with VLs represented continuously. Lower VL with sustained ART use was associated with a reduced probability of HIV transmission per act. Other factors modifying the HIV transmission probability per act included PrEP use, condom use, sexual position, circumcision, and a prevalent STI.

For HIV services, we represented an integrated HIV continuum of antiretroviral-based prevention and care, with HIV screening as the gateway to both.^18^ MSM engaged in HIV screening at regular intervals, calibrated to local surveillance data on the proportion of MSM with HIV who were diagnosed.^19^ MSM screening HIV-positive could then enter the HIV care continuum (linkage and retention in ART) while MSM who screened negative could enter the HIV prevention continuum (PrEP initiation, adherence, and persistence). MSM were linked to ART, and could cycle off and back on ART based on rates calibrated to local surveillance of care entry and VL suppression.^19^ MSM on ART achieved suppression after 3 months, with a rebound to set-point VL after halting ART.

The HIV prevention continuum consisted of initiation, adherence, and persistence in PrEP care for daily oral tenofovir/emtricitabine.^20^ MSM who tested HIV-negative and met indications for PrEP based on CDC guidelines were eligible to start.^21^ Eligible MSM then started PrEP based on an initiation probability generating a coverage level of 15%, consistent with Atlanta estimates.^22^ Heterogeneous PrEP adherence was modeled, with 78.4% meeting a high-adherence level that resulted in a 99% relative reduction in HIV acquisition risk.^23^ PrEP discontinuation was based on secondary estimates of the proportion of MSM who were retained in PrEP care at 6 months (57%).^24^ PrEP care consisted of routine HIV and STI screening.

Gonorrhea and chlamydia transmission were simulated along the same partnership network as HIV, but with disease recovery through either natural clearance or antibiotic treatment.^25^ STI transmission was directional and site-specific during anal intercourse at the rectal and urogenital sites. Men could be infected at both anatomical sites and with both gonorrhea and chlamydia. The symptomatic status of the newly acquired infections depended on site of infection, with most rectal infections asymptomatic and most urethral infections symptomatic.^26^ STI symptoms influenced the probability of testing and treatment, which reduced mean time to clearance. Successful treatment for an STI at one site resulted in clearance at the other site for dual-site infections.

### COVID-Related Impact on Behavior and Services

Experimental scenarios implemented reductions to the partnership network structure and HIV/STI service utilization individually and jointly. All scenarios were simulated for a period of 5 years in weekly time steps. A base scenario kept all parameters constant over that simulation window. Experimental scenarios first simulated one year of no change, followed by the initiation of either sexual distancing only, service interruption only, or both combined. In primary scenarios, service interruption lasted for 18 months and sexual distancing either for 18 months or 3 months. The 18-month window was selected based on predictions of the timeline for COVID-19 vaccine deployment; the 3-month distancing window was selected based on empirical data suggesting a more rapid behavioral rebound of sexual partnerships as early as June 2020.^9,10^ Sensitivity analyses varied the duration of both.

Distancing was modeled to varying levels by reducing network degree for casual partnerships and incidence for one-time partners; degree for main partnerships remained unchanged given minimal expected impact on cohabitating partners. Service interruption was reflected in four types of HIV/STI interventions: HIV screening, HIV PrEP (reduction in new users and increase in discontinuation for current users), HIV ART (through retention in care), and linked bacterial STI screening and treatment. These were reduced on a relative scale individually and jointly. We selected the joint 50% relative reduction in sexual behavior and clinical services as a key scenario to highlight in figures based on empirical data suggesting upwards of this level of change.^7,9^

### Calibration, Simulation, and Analysis

Given uncertainty in some model parameters, we calibrated the model with a Bayesian approach that defined prior distributions for these parameters and fit the model to empirical surveillance-based estimates of diagnosed HIV, NG, and CT incidence for the target population. This involved projecting incidence estimates to 2019 based on historical data and rising trends in cases over the past decade. After calibration, for each study scenario, we simulated the model 500 times and summarized the distribution of results with medians and 95% simulation intervals.

Three primary outcomes were: 1) standardized HIV and STI incidence per 100 person-years at risk (PYAR) at 2.5 years (or 18 months after start of COVID-related response); 2) standardized cumulative incidence over 5 years per 1000 disease-susceptible MSM; and 3) the total 5-year population impact, which was calculated in two steps. We first multiplied the standardized cumulative incidence by estimates of the total susceptible population size of MSM in the Atlanta metropolitan area (102,642 sexually active MSM for STI outcomes, and 87,723 sexually active HIV-negative MSM for HIV outcomes^27^) to quantify the total population 5-year incidence. We then subtracted this total population incidence for each scenario from the value in the base scenario to obtain an absolute difference.

## RESULTS

**Figure 1** visualizes the primary scenarios of 18 months of service interruption and 18 months of sexual distancing. The panels show standardized incidence rates of HIV and STIs for scenarios in which sexual behavior and services were jointly reduced by 50% during the COVID period. **Panel A** shows that distancing in the absence of service reduction (green line) was associated with a decrease in HIV incidence, whereas service reduction only (red line) was associated with an increase in HIV incidence. HIV incidence changes in both scenarios persisted after the resumption of baseline behavior and services at year 2.5 (week 130). In the combined scenario (blue line), these relative changes in behavior and services effectively largely each other, resulting in minimal difference in HIV incidence compared with the base (no change) scenario. **Panel B** shows the impact of the same scenarios on combined gonorrhea and chlamydia incidence. Sexual distancing had a strong and sustained reduction in STI incidence. With only service interruption but no distancing, STI incidence increased substantially. In the combined scenario, the projected incidence was slightly lower initially, rebound after sexual distancing ends, and then continued to decline again through year 5 (week 260).

**Figure 1.**
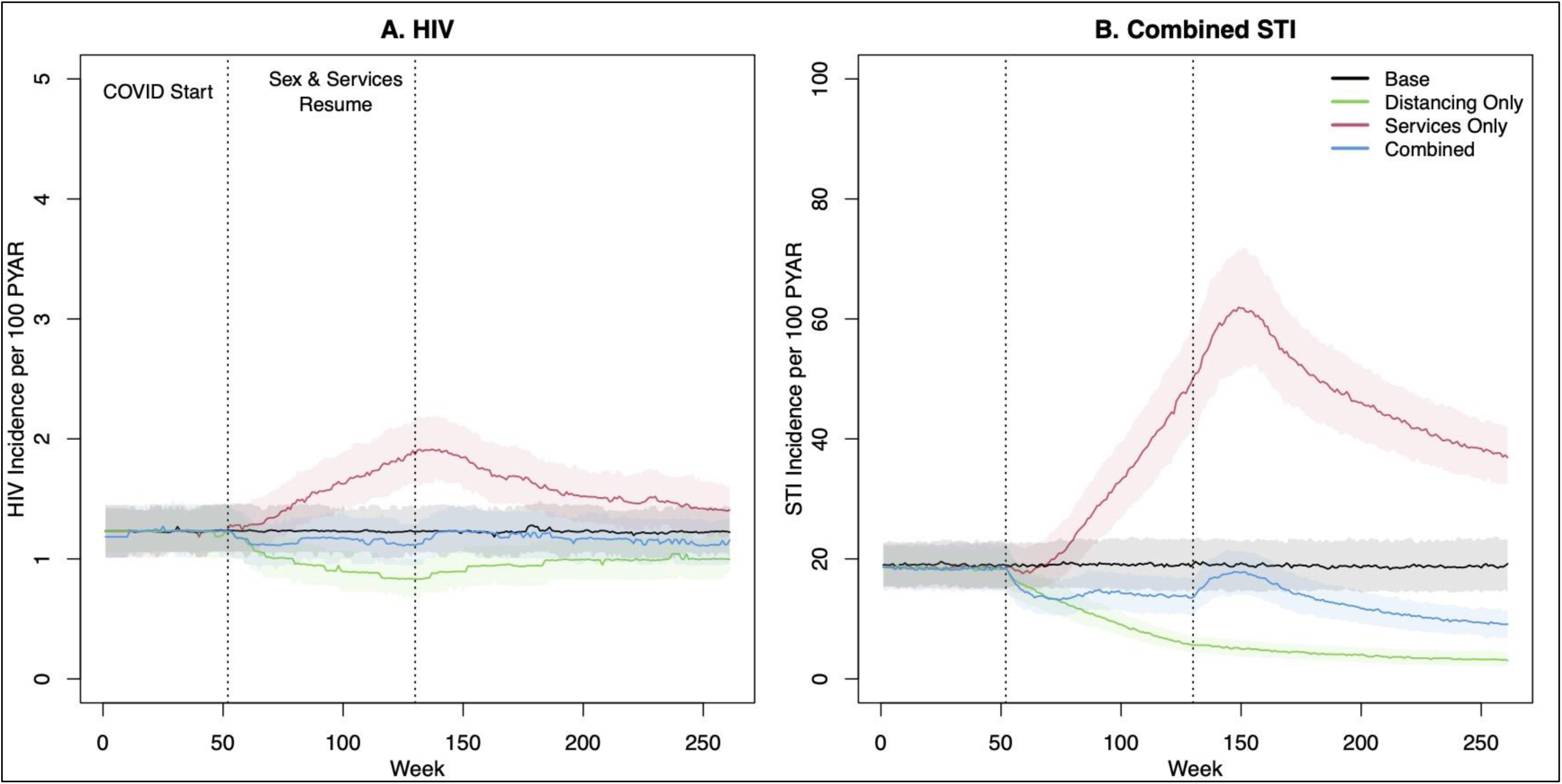
HIV and combined STI incidence before, during, and after an 18-month period of clinical service interruption and 18-month period of sexual distancing. Model scenarios within each panel compare a 50% relative reduction in behavior only, service interruption only, or both jointly against the base (no change) model. Thick lines show median values and bands show inter-quartile range of values across 500 simulations per scenario.

**Table 1** quantifies the estimated impact of the sexual-distancing-only scenarios with an 18-month duration. A relative 50% reduction in casual and one-time networks jointly (corresponding to the green line scenario in Figure 1) resulted in a decrease in HIV incidence from 1.23 per 100 PYAR to 0.79, a 36% relative reduction. The 5-year cumulative incidence per 1,000 susceptible MSM was 51.4 cases in that scenario, compared to 62.3 cases in the base scenario, corresponding with a 17% reduction in HIV incidence. The impact on the point incidence at year 2.5 was more extreme than the impact on the cumulative incidence across 5 years because the point incidence rebounded to the baseline level after distancing and service interruptions ended. The total population impact on HIV incidence for Atlanta MSM was projected in that scenario to be −966 cases, or 193 fewer cases per year, compared to the base scenario. The relative size of reductions in casual partnerships correlated with stronger relative declines in HIV incidence than comparable relative reductions in one-time partnerships. This was driven by the underlying behavioral patterns, in which coital frequency was higher and condom use lower for casual partnerships. The mechanistic impact on network degree statistics is provided in **Supplemental Table 14**.

**Table 1.** Changes in HIV and Combined Gonorrhea & Chlamydia Incidence Following an 18-Month Period of Sexual Distancing, with no Associated Changes in HIV/STI Prevention or Treatment Service Utilization

| Scenario | HIV |  |  | Combined STI |  |  |
| --- | --- | --- | --- | --- | --- | --- |
|  | *Incidence Rate<sup>A</sup>*<br>(95% SI) | *Cumul. Incidence<sup>B</sup>*<br>(95% SI) | *Population Impact<sup>C</sup>*<br>(95% SI) | *Incidence Rate<sup>A</sup>*<br>(95% SI) | *Cumul. Incidence<sup>B</sup>*<br>(95% SI) | *Population Impact<sup>C</sup>*<br>in Thousands (95% SI) |
| **Base Scenario** |  |  |  |  |  |  |
| No Changes | 1.23 (0.56, 2.05) | 62.3 (54.5, 70.7) | – | 19.39 (8.89, 31.70) | 960.4 (477.0, 1461.6) | – |
| **Reduction in Casual Network Degree & One-Time Rate** |  |  |  |  |  |  |
| 25% | 1.00 (0.44, 1.77) | 56.9 (49.8, 64.2) | -488 (-518, -459) | 10.67 (4.76, 18.55) | 644.1 (330.8, 1033.4) | -31.4 (-33.4, -29.3) |
| 50% | 0.79 (0.33, 1.56) | 51.4 (45.1, 58.0) | -966 (-995, -939) | 5.85 (2.40, 9.76) | 445.9 (227.8, 710.9) | -51.8 (-53.5, -50.3) |
| 90% | 0.66 (0.11, 1.32) | 44.5 (37.3, 171.0) | -1547 (-1579, -1515) | 1.30 (0.51, 2.73) | 291.4 (154.2, 1409.6) | -67.7 (-69.1, -66.3) |
| **Reduction in Casual Network Degree** |  |  |  |  |  |  |
| 25% | 1.10 (0.44, 1.90) | 58.2 (50.2, 65.5) | -368 (-397, -340) | 14.33 (6.86, 24.59) | 728.1 (384.7, 1186.3) | -22.2 (-24.2, -20.2) |
| 50% | 0.90 (0.33, 1.68) | 53.2 (46.7, 61.3) | -781 (-812, -752) | 10.54 (5.01, 18.05) | 593.7 (332.6, 945.5) | -37.1 (-39.0, -35.3) |
| 90% | 0.67 (0.22, 1.45) | 46.3 (38.8, 222.4) | -1381 (-1417, -1348) | 2.69 (1.04, 5.04) | 341.3 (180.4, 1948.7) | -62.4 (-64.2, -60.9) |
| **Reduction in One-Time Rate** |  |  |  |  |  |  |
| 25% | 1.12 (0.45, 2.01) | 60.5 (53.1, 68.8) | -169 (-199, -141) | 13.69 (6.24, 22.20) | 783.4 (400.1, 1186.9) | -17.7 (-19.8, -15.6) |
| 50% | 1.11 (0.44, 1.88) | 58.7 (51.6, 67.1) | -322 (-352, -292) | 9.52 (3.71, 15.84) | 655.2 (298.8, 1028.8) | -30.9 (-32.9, -28.9) |
| 90% | 0.89 (0.33, 1.57) | 55.9 (49.5, 63.6) | -561 (-588, -532) | 4.90 (2.07, 9.12) | 470.8 (223.1, 785.3) | -49.7 (-51.4, -48.0) |

<sup>A</sup> Standardized rate per 100 person-years at risk at 2.5 years
<sup>B</sup> Standardized cumulative incidence over 5 years per 1000 susceptible MSM
<sup>C</sup> Difference, compared to base scenario, in 5-year cumulative incidence for total susceptible population of MSM in Atlanta

The patterns were generally similar for STI incidence across the same **Table 1** scenarios of sexual distancing only with an 18-month duration. The baseline STI incidence rate was 19.39 (per 100 PYAR), but 50% joint network distancing resulted in an estimated incidence of 5.85 at year 2.5 (a 70% reduction) and a cumulative incidence of 446 cases compared to 960 in the base scenario (a 54% reduction). The 5-year population impact of this scenario was a reduction in total STI cases by over 50,000. Also compared to HIV, 50% reductions within each partnership sub-network were associated with similar declines in STI incidence, whereas HIV was more strongly impacted by casual network reductions. Individual STI outcomes are provided in **Supplemental Table 15**.

**Table 2** quantifies the impact of HIV/STI service reductions in the absence of sexual distancing. The scenario in which all services were reduced by 50% corresponds to the red lines in the **Figure 1**. Reductions in PrEP had a more moderate impact on HIV incidence than reductions in ART. This was because baseline PrEP coverage was relatively low (15%) in Atlanta, and COVID-related changes only applied to the PrEP-indicated population. In comparison, changes in ART retention would impact the care of the entire HIV-diagnosed population, subsequently affecting transmission through lower levels of VL suppression. For STIs, reductions in STI treatment had a dramatic impact on STI incidence, which was held in equilibrium in the base scenario by high levels of routine screening. Gaps in STI screening, which were projected to dramatically increase STI incidence, also had downstream effects on HIV incidence through the biological relationship between prevalent STI infection and HIV acquisition risk. Process outcomes associated for these scenarios are provided in **Supplemental Table 16** and individual STI outcomes in **Supplemental Table 17**.

**Table 2.** Changes in HIV and Combined Gonorrhea & Chlamydia Incidence Following an 18-Month Period of Reduced HIV/STI Prevention or Treatment Service Utilization, with no Associated Changes in Behavior

| Scenario | HIV |  |  | Combined STI |  |  |
| --- | --- | --- | --- | --- | --- | --- |
|  | *Incidence Rate<sup>A</sup>* | *Cumul. Incidence<sup>B</sup>* | *Population Impact<sup>C</sup>* | *Incidence Rate<sup>A</sup>* | *Cumul. Incidence<sup>B</sup>* | *Population Impact<sup>C</sup>* |
|  | (95% SI) | (95% SI) | (95% SI) | (95% SI) | (95% SI) | in Thousands (95% SI) |
| **Base Scenario** |  |  |  |  |  |  |
| No Changes | 1.23 (0.56, 2.05) | 62.3 (54.5, 70.7) | – | 19.39 (8.89, 31.70) | 960.4 (477.0, 1461.6) | – |
| **Reduction in All Services** |  |  |  |  |  |  |
| 25% | 1.56 (0.78, 2.44) | 68.7 (60.4, 78.0) | 558 (527, 590) | 35.52 (17.93, 52.41) | 1481.5 (786.4, 2081.4) | 52.0 (49.5, 54.5) |
| 50% | 1.90 (1.01, 3.02) | 76.9 (67.0, 86.5) | 1272 (1237, 1306) | 48.83 (27.53, 71.19) | 1862.1 (1114.8, 2636.8) | 92.3 (89.7, 94.9) |
| 90% | 3.26 (2.02, 4.63) | 105.6 (94.7, 118.3) | 3785 (3748, 3822) | 64.23 (38.05, 90.76) | 2261.8 (1475.7, 3045.5) | 132.7 (130.1, 135.3) |
| **Reduction in PrEP Initiation** |  |  |  |  |  |  |
| 25% | 1.24 (0.67, 2.14) | 63.2 (55.3, 71.6) | 63 (32, 95) | 18.47 (8.73, 31.62) | 943.7 (507.9, 1502.9) | -1.0 (-3.3, 1.5) |
| 50% | 1.33 (0.56, 2.13) | 63.9 (55.9, 72.0) | 131 (101, 165) | 18.50 (9.23, 31.68) | 936.4 (489.0, 1469.6) | -1.5 (-3.7, 0.7) |
| 90% | 1.35 (0.67, 2.25) | 65.2 (57.5, 74.4) | 261 (228, 290) | 19.50 (8.98, 32.18) | 991.6 (515.9, 1487.5) | 2.3 (0.1, 4.6) |
| **Reduction in HIV Screening** |  |  |  |  |  |  |
| 25% | 1.24 (0.56, 2.13) | 63.2 (54.9, 71.4) | 78 (47, 109) | 18.61 (8.98, 31.28) | 946.1 (485.3, 1462.1) | -2.2 (-4.4, 0.0) |
| 50% | 1.33 (0.56, 2.14) | 64.3 (56.2, 73.2) | 176 (144, 209) | 18.58 (8.86, 30.76) | 927.0 (496.8, 1476.8) | -1.4 (-3.5, 0.7) |
| 90% | 1.45 (0.67, 2.35) | 66.3 (58.0, 75.9) | 350 (319, 384) | 18.79 (9.40, 28.83) | 951.0 (495.8, 1424.5) | -0.6 (-2.6, 1.6) |
| **Reduction in ART Retention** |  |  |  |  |  |  |
| 25% | 1.24 (0.56, 2.24) | 64.0 (56.9, 72.7) | 157 (124, 189) | 18.71 (8.72, 30.16) | 943.7 (475.9, 1425.8) | -1.5 (-3.4, 0.6) |
| 50% | 1.45 (0.67, 2.48) | 67.8 (60.0, 76.1) | 469 (437, 498) | 18.51 (8.77, 29.76) | 943.0 (451.9, 1430.1) | -1.7 (-4.0, 0.6) |
| 90% | 2.26 (1.24, 3.30) | 89.0 (79.0, 97.7) | 2324 (2291, 2357) | 18.21 (9.20, 29.63) | 932.0 (508.5, 1404.8) | -2.4 (-4.5, -0.2) |
| **Reduction in NG/CT Treatment** |  |  |  |  |  |  |
| 25% | 1.24 (0.66, 2.26) | 64.5 (56.2, 73.2) | 184 (155, 217) | 35.31 (17.10, 54.45) | 1459.3 (768.0, 2127.0) | 51.2 (48.5, 54.0) |
| 50% | 1.44 (0.67, 2.35) | 66.3 (57.7, 75.9) | 346 (311, 377) | 49.38 (27.71, 71.72) | 1851.2 (1172.2, 2568.7) | 92.6 (89.9, 95.3) |
| 90% | 1.47 (0.77, 2.47) | 68.7 (60.6, 78.0) | 569 (536, 603) | 66.62 (41.00, 90.51) | 2310.7 (1562.6, 3010.2) | 137.9 (134.8, 140.9) |

<sup>A</sup> Standardized rate per 100 person-years at risk at 2.5 years
<sup>B</sup> Standardized cumulative incidence over 5 years per 1000 susceptible MSM
<sup>C</sup> Difference, compared to base scenario, in 5-year cumulative incidence for total susceptible population of MSM in Atlanta

The top half of **Table 3** shows the impact of combined sexual distancing and service reduction for 18 months. The 50% network reduction and 50% service reduction scenario corresponds to the blue lines in **Figure 1**. For HIV, commensurate relative reductions in behavior and services by the same amounts generally kept incidence similar to the base scenario. The population impact of paired scaled-down scenarios had a minor protective effect at lower reductions (25%/25%: −69; 50%/50%: −227) but resulted in slightly higher incidence for the most extreme reductions (90%/90%: 215). For STIs, in contrast, declines in behavior strongly overwhelmed the declines in services, with a net reduction of 23,800 cases in the 50%/50% paired scaled-down scenario.

**Table 3.** Changes in HIV and Combined Gonorrhea & Chlamydia Incidence Following Joint Behavioral Change and Reduced HIV/STI Prevention and Treatment Service Utilization

| Scenario | HIV |  |  | Combined STI |  |  |
| --- | --- | --- | --- | --- | --- | --- |
|  | *Incidence Rate<sup>A</sup>* | *Cumul. Incidence<sup>B</sup>* | *Population Impact<sup>C</sup>* | *Incidence Rate<sup>A</sup>* | *Cumul. Incidence<sup>B</sup>* | *Population Impact<sup>C</sup>* |
|  | (95% SI) | (95% SI) | (95% SI) | (95% SI) | (95% SI) | in Thousands (95% SI) |
| **Base Scenario** |  |  |  |  |  |  |
| No Changes | 1.23 (0.56, 2.05) | 62.3 (54.5, 70.7) | – | 19.39 (8.89, 31.70) | 960.4 (477.0, 1461.6) | – |
| **Sexual Distancing for 18 Months, Service Reduction for 18 Months** |  |  |  |  |  |  |
| Reduced Casual/One-Time Networks by 25% |  |  |  |  |  |  |
| Services by -25% | 1.22 (0.55, 2.13) | 61.7 (54.1, 69.1) | -69 (-98, -38) | 20.01 (9.89, 30.90) | 955.4 (480.9, 1416.9) | 0.0 (-2.3, 2.1) |
| Services by -50% | 1.46 (0.68, 2.36) | 67.6 (59.7, 76.2) | 465 (435, 496) | 27.45 (14.61, 43.32) | 1198.4 (704.1, 1750.8) | 25.1 (22.8, 27.5) |
| Services by -90% | 2.49 (1.48, 3.71) | 92.0 (82.9, 102.5) | 2625 (2590, 2661) | 35.71 (20.44, 55.03) | 1470.9 (869.8, 2089.8) | 51.9 (49.4, 54.5) |
| Reduced Casual/One-Time Networks by 50% |  |  |  |  |  |  |
| Services by -25% | 0.90 (0.33, 1.67) | 54.5 (47.8, 61.6) | -689 (-718, -660) | 10.49 (4.95, 17.57) | 611.5 (313.6, 977.1) | -35.4 (-37.1, -33.7) |
| Services by -50% | 1.12 (0.55, 2.01) | 59.9 (52.1, 66.8) | -227 (-257, -198) | 13.59 (7.16, 22.00) | 736.8 (375.3, 1135.6) | -23.8 (-25.8, -21.9) |
| Services by -90% | 1.90 (1.11, 2.84) | 79.9 (71.5, 88.4) | 1532 (1500, 1565) | 17.38 (9.52, 27.69) | 860.3 (511.8, 1323.4) | -9.3 (-11.3, -7.2) |
| Reduced Casual/One-Time Networks by 90% |  |  |  |  |  |  |
| Services by -25% | 0.66 (0.11, 1.44) | 47.2 (39.0, 188.9) | -1298 (-1331, -1262) | 2.42 (1.03, 4.39) | 318.1 (167.5, 1232.6) | -64.7 (-66.1, -63.1) |
| Services by -50% | 0.78 (0.33, 1.68) | 49.8 (41.9, 348.0) | -1074 (-1107, -1040) | 3.11 (1.30, 5.28) | 333.8 (183.7, 3540.3) | -62.8 (-64.4, -61.3) |
| Services by -90% | 1.33 (0.56, 2.34) | 64.5 (55.3, 384.4) | 215 (178, 256) | 3.63 (1.66, 6.20) | 350.3 (191.6, 5041.2) | -60.5 (-62.2, -59.0) |
| **Sexual Distancing for 3 Months, Service Reduction for 18 Months** |  |  |  |  |  |  |
| Reduced Casual/One-Time Networks by 25% |  |  |  |  |  |  |
| Services by -25% | 1.45 (0.67, 2.47) | 67.4 (58.6, 77.0) | 423 (388, 455) | 31.06 (16.36, 49.10) | 1302.8 (741.7, 2008.1) | 37.2 (34.7, 39.9) |
| Services by -50% | 1.80 (0.89, 2.92) | 74.8 (65.4, 84.4) | 1074 (1039, 1108) | 44.10 (22.59, 64.69) | 1719.4 (953.5, 2348.6) | 76.6 (74.0, 79.2) |
| Services by -90% | 3.16 (1.93, 4.54) | 103.1 (91.9, 114.2) | 3559 (3523, 3596) | 58.60 (31.00, 81.66) | 2105.9 (1273.2, 2829.7) | 116.4 (113.7, 119.4) |
| Reduced Casual/One-Time Networks by 50% |  |  |  |  |  |  |
| Services by -25% | 1.45 (0.67, 2.25) | 65.3 (56.9, 74.3) | 252 (220, 283) | 27.08 (13.90, 43.78) | 1184.6 (640.5, 1829.3) | 23.7 (21.2, 26.0) |
| Services by -50% | 1.78 (0.89, 2.81) | 72.6 (63.0, 82.0) | 890 (858, 923) | 38.45 (19.20, 55.92) | 1529.3 (857.3, 2106.2) | 57.5 (55.1, 60.0) |
| Services by -90% | 2.93 (1.70, 4.39) | 99.2 (88.0, 111.0) | 3240 (3202, 3277) | 50.23 (28.22, 71.85) | 1871.4 (1141.2, 2505.3) | 93.1 (90.5, 95.8) |
| Reduced Casual/One-Time Networks by 90% |  |  |  |  |  |  |
| Services by -25% | 1.13 (0.55, 1.99) | 56.5 (49.9, 64.2) | -513 (-543, -482) | 11.79 (5.30, 20.57) | 613.4 (320.9, 972.0) | -34.9 (-36.6, -33.0) |
| Services by -50% | 1.34 (0.67, 2.23) | 61.9 (54.4, 69.6) | -46 (-75, -18) | 17.15 (8.05, 27.05) | 789.0 (384.4, 1181.6) | -17.7 (-19.6, -15.9) |
| *Services by -90%* | 2.27 (1.34, 3.35) | 83.2 (75.3, 92.7) | 1835 (1803, 1867) | 22.59 (11.02, 35.04) | 954.1 (518.1, 1444.7) | 0.8 (-1.4, 2.9) |

<sup>A</sup> Standardized rate per 100 person-years at risk at 2.5 years
<sup>B</sup> Standardized cumulative incidence over 5 years per 1000 person-years at risk
<sup>C</sup> Difference, compared to base scenario, in 5-year cumulative incidence for total susceptible population of MSM in Atlanta

**Figure 2** demonstrates how the interaction of the duration of sexual distancing and service interruptions impacts the standardized cumulative incidence outcomes. Similar to **Figure 1**, the scenarios here reflected a 50% relative reduction in both behavior and services during the eligible change period. HIV and STI incidence were lowest (53 and 533 cases per 1,000 susceptible, respectively) when service interruption lasted for 3 months and distancing lasted for 18 months. Both HIV and combined STI incidence were highest (73 and 1,529 cases, respectively) when services were interrupted for 18 months but sexual distancing occurred for 3 months.

**Figure 2.**
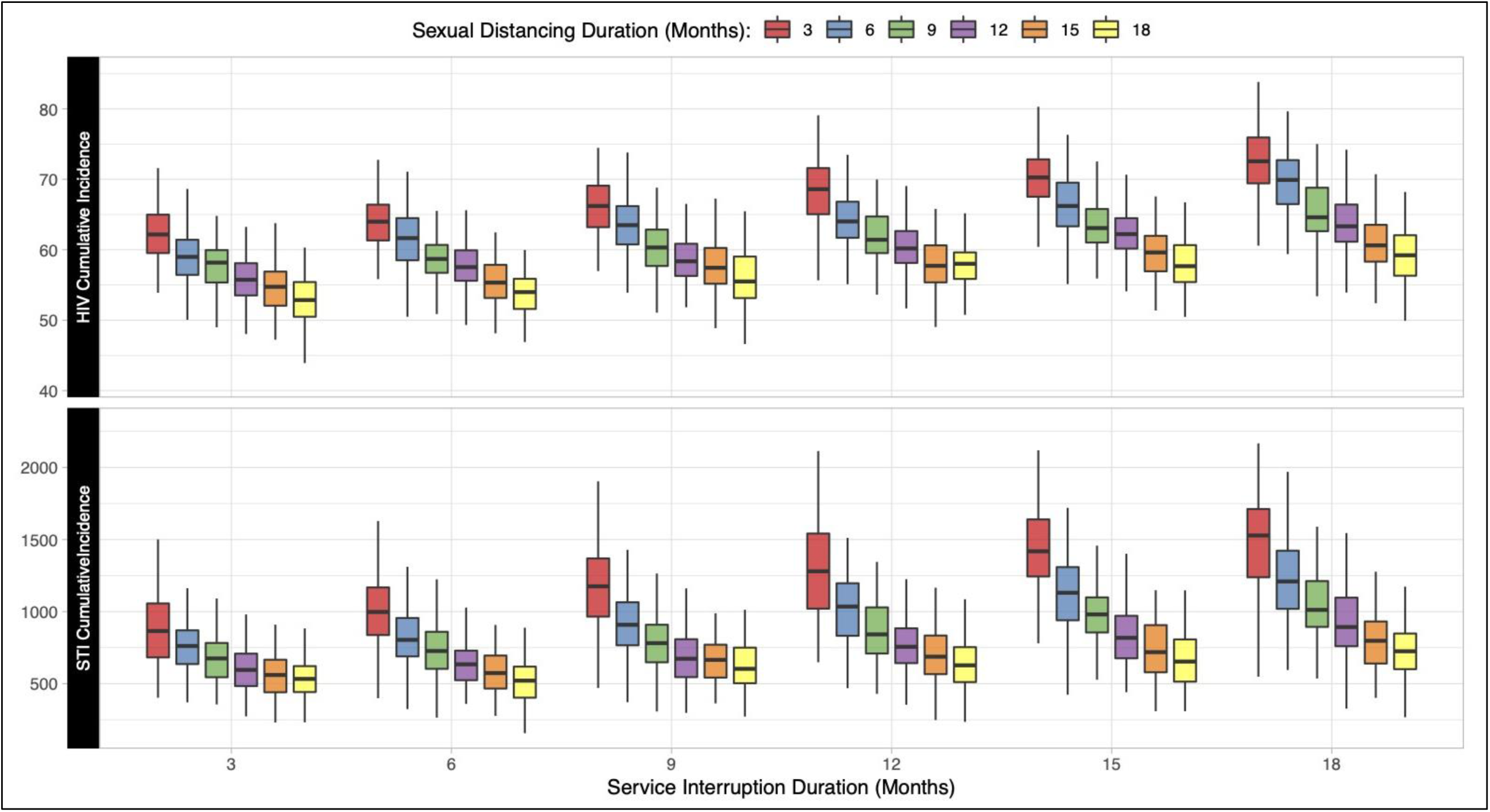
Relationship between the duration of sexual distancing and service interruption on cumulative (5-year) incidence of HIV and STIs per 1000 susceptible MSM. Individual box and whiskers display the distribution of cumulative incidence across 500 simulations within each scenario.

Finally, we explored that last scenario further in **Figure 3** and the bottom of **Table 3**. Here the resumption time for sexual behavior was varied independently from the resumption time for services. For HIV, 3-month distancing in the absence of service change had no substantive impact on the trajectory of HIV incidence (green line), and therefore was unable to counterbalance the effects of service interruption in the combined scenario (blue line). Higher point incidence lasted through the end of 5 years. Over 5 years, this resulted in 890 excess HIV cases. For STIs, incidence in the combined scenario followed a similar pattern as for HIV, but to a more extreme level. STI incidence in the combined scenario (blue line) more closely tracked the scenario with service interruption only (red line), and STI incidence did not return to baseline values through 5 years. This resulted in an excess of 57,500 cases over 5 years.

**Figure 3.**
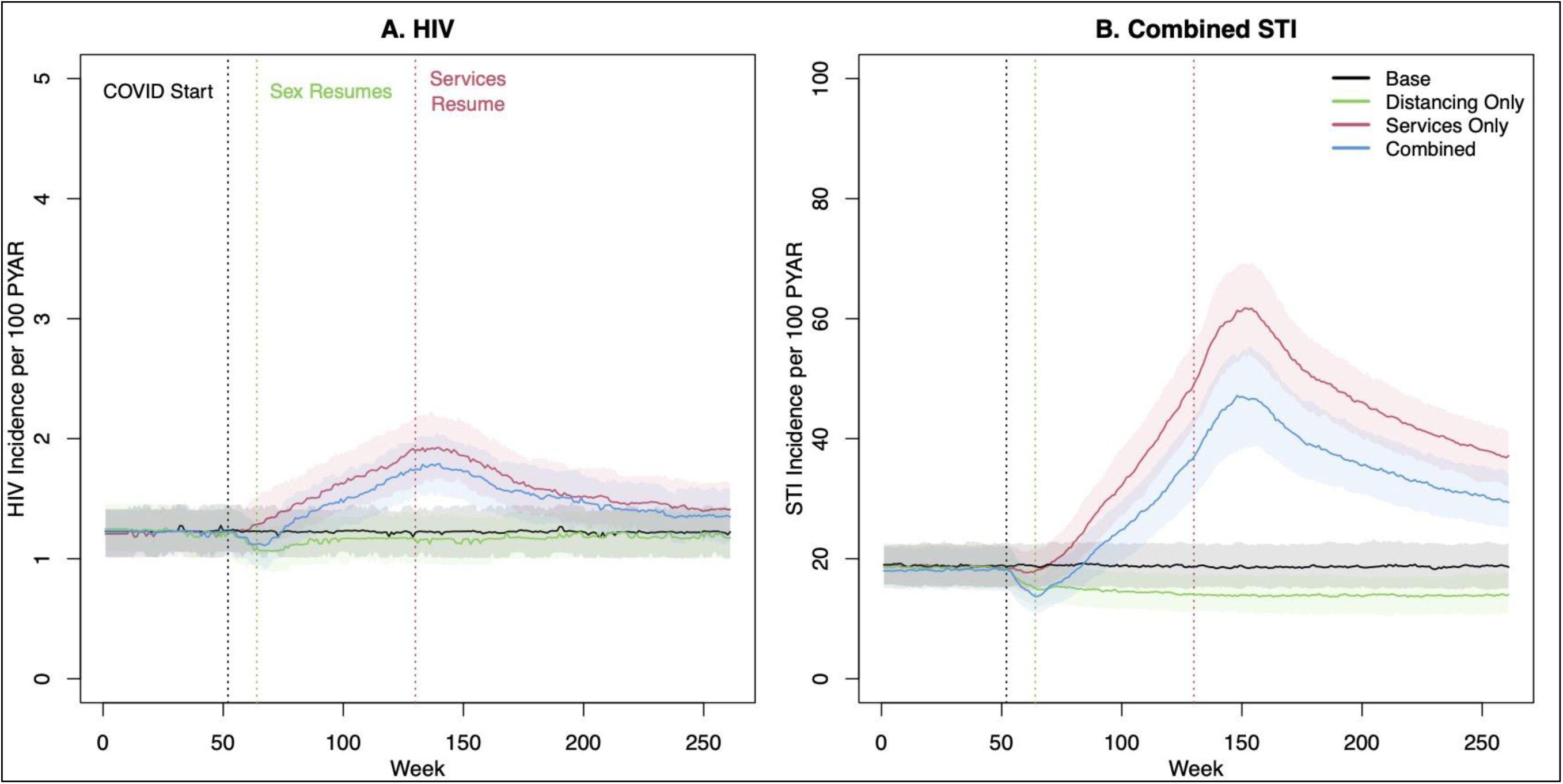
HIV and combined STI incidence before, during, and after an 18-month period of clinical service interruption and 3-month period of sexual distancing. Model scenarios within each panel compare a 50% relative reduction in behavior only, service interruption only, or both jointly against the base (no change) model. Thick lines show median values and bands show inter-quartile range of values across 500 simulations per scenario.

## DISCUSSION

This study projected the 5-year impact of COVID-related sexual behavior changes (“sexual distancing”) and interruption of clinical services on the incidence of HIV, gonorrhea, and chlamydia among MSM in the Atlanta area. We found that the magnitude and timing of epidemiological impact depended on the infection and on the relative extent and durability of the COVID-related changes. Durable sexual distancing could offset (for HIV) or overcome (for STIs) the excess incidence attributable to an equal period of clinical service interruption. However, sexual activity rebounding while service interruption persists would lead to higher incidence for both HIV and STIs. Based on current estimates of behavioral and clinical change, and future predictions of an 18-month clinical service interruption duration, we project an excess of nearly 900 HIV cases and over 57,000 STI cases just among Atlanta MSM over the next 5 years. Our findings suggest that immediate action is needed to address the indirect effects of the COVID-19 pandemic on the HIV/STI epidemic.

The protective effects of sexual distancing on both HIV and STI incidence highlight the need to understand and address the extent and durability of behavior change during the COVID-19 pandemic. In our model, sexual partner reduction had a protective effect on HIV and STI incidence. Empirical data suggests a wide heterogeneity of changes in sexual activity during initial COVID restrictions in March 2020.^7,8^ However, fewer studies have characterized the timing of behavioral rebounds; some have suggested a return to pre-COVID levels starting as early as June 2020.^9,10^ Our model suggests that such a transient change will have no substantive impact for HIV and only a minor impact for STIs. The epidemiological impact of behavioral responses to COVID-19 highlight the need for effective sexual health messaging that supports sustainable behavior changes throughout the pandemic, which could have a protective effect on both HIV/STIs and SARS-CoV-2 transmission. Local health departments, such as the New York City Department of Health, have developed such guidelines.^28^ Sexual health messaging should adopt a harm reduction framework.^29^

The projected detrimental effects on HIV and STI incidence of clinical service interruption in the model demonstrate the critical importance of maintaining sexual health services amidst the COVID-19 pandemic response. Our model evaluated four clinical services for which there was already evidence of interruption to the relative degree in our scenarios.^7^ In some jurisdictions, health department staff assigned to HIV/STI partner services have been redeployed for COVID contact tracing.^13^ Interruption of ART care for persons living with HIV had the largest impact on projected excess HIV incidence in our model. This finding is consistent with two other models of the impact of COVID-19 on HIV outcomes, focused on low/middle-income countries.^1,30^ Minimizing service interruption will require innovative approaches to ensure access to clinical services and overcome common barriers to care during the COVID pandemic, including travel limitations and gaps in health insurance. These approaches will remain important even as sexual health services return to pre-COVID capacity and long-lasting impacts on health care access affect re-engagement in services.

### Limitations

The primary limitation of our model projections is the future uncertainty about the types and durability of both sexual distancing and service reduction. While our sensitivity analysis (**Figure 2**) explored durability, both components may change or rebound in different ways from those modeled. Modeling changes in the COVID-19 era required mapping data on broader aggregate reductions onto individual rate-based model parameters. Future empirical research on sexual distancing and service interruption should measure these changes with more individual-level specificity within and across persons. Second, we assumed that there was no correlation between changes in individual behavior and changes in service engagement. This decision was based on a re-analysis of one published study,^7^ provided in **Supplemental Table 4**.**1**, showing limited evidence of such individual-level correlation between change in sexual partner numbers and change in service access (small correlations were observed for HIV and STI screening only). Additionally, some group-level age-related changes in both behavior and services have been noted, but we did not include that heterogeneity in the model because it did not emerge in individual-level correlations. Third, our model did not explicitly represent the transmission dynamics of COVID-19 that may result in changes to sexual networks based on real or perceived COVID risk (e.g., partner selection by COVID status or risk factors, or other disruptions to travel to meet sex partners that could result in concentrated transmission clusters). This is an important topic for future data collection and modeling studies. Finally, our target population was MSM in the Atlanta area, a population with lower baseline access to HIV/STI services and a higher baseline of infection than other populations and areas. Therefore, our standardized results may not be scalable to populations with a different epidemiological context.

### Conclusions

The global COVID-19 pandemic could present substantial challenges to the prevention and control of other infectious diseases, including HIV and STIs, as a result of clinical service interruptions. While sexual behavior change may offset service interruptions, this will depend on overlap in the timing of these changes. With transient behavior change but persistent service interruptions, we project major increases in HIV and STI incidence that may take years to return to pre-COVID levels. This calls for improved sexual health messaging and innovative approaches to addressing service gaps during the ongoing COVID-19 pandemic.

## Data Availability

Model code and data are available at the Github repositories linked in the Supplemental Appendix document.

## Notes

**FUNDING** This work was supported by grants from National Institutes of Health grant (R01 AI138783 and P30 AI050409) and the MAC AIDS Fund. JLM is supported in part by the National Institute of Allergy and Infectious Diseases (grant K01 AI122853).

### Competing Interest Statement

JLM has previously consulted for Kaiser Permanente Northern California on a research grant from Gilead Sciences. DW has consulted for Sanofi-Pasteur on unrelated topics.

### Funding Statement

This work was supported by grants from National Institutes of Health grant (R01 AI138783 and P30AI050409) and the MAC AIDS Fund. JLM is supported in part by the National Institute of Allergy and Infectious Diseases (grant K01 AI122853).

### Author Declarations

Emory University Institutional Review Board approved the human subjects research component of the ARTnet and AMIS studies that generated primary data for use in this model.

### Summary of Updates

Edited text throughout for clarity. Fixed plotting of temporal outcomes in Figures 1 and 3.

